# Systematic review reveals multiple sexually antagonistic polymorphisms affecting human disease and complex traits

**DOI:** 10.1101/2020.12.16.20248300

**Authors:** Jon Alexander Harper, Tim Janicke, Edward H. Morrow

## Abstract

An evolutionary model for sex differences in disease risk posits that alleles conferring higher risk in one sex may be protective in the other. These sexually antagonistic (SA) alleles are predicted to be maintained at frequencies higher than expected under purifying selection against unconditionally deleterious alleles, but there are apparently no examples in humans. Discipline-specific terminology, rather than a genuine lack of such alleles, could explain this disparity. We undertook a two-stage review of evidence for SA polymorphisms in humans using search terms from (i) evolutionary biology and (ii) biomedicine. While the first stage returned no eligible studies, the second revealed 51 genes with sex-opposite effects, 22 increased disease risk or severity in one sex but protected the other. Those with net positive effects occurred at higher frequencies. None were referred to as SA. Our review reveals significant communication barriers to fields as a result of discipline-specific terminology.

## Introduction

In species with separate sexes an evolutionary conflict at the level of individual genetic loci can occur, where alleles that are favoured by selection in one sex are selected against in the other (i.e intralocus sexual conflict, Parker, 1979). This sexually antagonistic (SA) form of selection is thought to be driven by differences in how the two sexes maximize their fitness and concomitant unequal variances in reproductive success. Research into SA selection has expanded recently with the recognition that it feeds into several important evolutionary processes. It is primarily thought to drive the evolution of sexual dimorphism and trait diversification (Lande, 1980; Pennell et al., 2016; Rice, 1984), including sex-biased gene expression (Ellegren and Parsch, 2007). As a potential driver of balancing selection, it has also been implicated in the maintenance of genetic variation (Connallon and Clark, 2013; Grieshop and Arnqvist, 2018) that would otherwise be eroded by directional selection. Most recently, it has been suggested that genetic variation at SA loci could contribute to the occurrence of a number of common human diseases (Morrow, 2015; Morrow and Connallon, 2013), which also show considerable variation between the sexes in terms of their prevalence, severity and age of onset (Ober et al., 2008; Rigby and Kulathinal, 2015).

Despite the interest in sexual antagonism as an evolutionary force, determining the identity of SA loci remains a major challenge (Ruzicka et al., 2020). An early empirical milestone in the field of sexual conflict was achieved by quantitative genetic studies demonstrating that genomes in a laboratory adapted population of *Drosophila melanogaster* harbour significant amounts of SA standing genetic variation (Chippindale et al., 2001; Rice, 1992). A number of other systems have shown similar results, albeit using different methods, including invertebrates and vertebrates from both lab and wild populations (Bonduriansky and Chenoweth, 2009; Foerster et al., 2007; Mills et al., 2011). With the advent of advanced genomic tools, two model systems report specific examples of SA genetic loci: in Atlantic salmon (*Salmo salar*) the VGLL3 locus (Barson et al., 2015), and in *D. melanogaster* the DDT-R locus (Rostant et al., 2015), the Ala-278-Thr polymorphism in the mitochondrial genome (Camus et al., 2015), and multiple candidate loci from a recent genome-wide association study (Ruzicka et al., 2019).

Although humans have also been shown to experience SA selection for some quantitative traits (Garver-Apgar et al., 2011; Stearns et al., 2012; Stulp et al., 2012), and there are numerous reports of genetic loci with sex-specific effects (i.e. effects that differ in magnitude between the sexes) on multiple complex traits or disease phenotypes (Gilks et al., 2014; Winkler et al., 2015), there are apparently no clear examples of SA loci in humans, which is inconsistent with theoretical expectations (Connallon and Clark, 2014). One potential explanation is that sexual antagonism is a weak selective force in humans, with sexual dimorphism being rather limited (Dixson, 2009; Short, 1979). This may indicate that concordant selection pressures between the sexes have dominated our evolutionary history. But the recent quantitative genetic studies challenge this view (Stearns et al., 2012; Stulp et al., 2012), and indeed theory predicts there is an inevitability to SA loci occurring in organisms with separate sexes (Connallon and Clark, 2014; Parker et al., 1972). Thus, there is a clear disparity between, on the one hand, theoretical expectations and quantitative genetic evidence that SA selection does occur in humans, and on the other hand, a complete lack of specific examples of SA loci. All this in the context of decades of research into how individual genetic variants influence disease profiles.

An alternative explanation for the absence of documented SA loci in humans is that since biomedical science does not use the same terminology for SA effects that evolutionary biology uses, there may in fact be examples that have been misclassified and therefore remain hidden in the literature. For example, genetic loci with sex-specific effects are frequently referred to as *sex-dependent* effects in the biomedical literature. Other terms are also used for related concepts, including *sex-different* effects. Furthermore, *sex-opposite* effects (i.e. a positive effect in one sex and a negative in the other) could be SA effects if the phenotype in question is deleterious, as disease phenotypes often are, since they can influence an individual’s survival or reproductive fitness. The biomedical literature also frequently uses the terms *sex* and *gender* interchangeably, with *males* and *females* more often than not referred to as *men and women*, or *boys and girls* (Khramtsova et al., 2019). There is therefore ample scope for discipline-specific terms to be used for describing the same concept, which may hinder communication between disciplines and obscure examples of SA loci. Moreover, although there has been some progress in introducing concepts from evolutionary biology into the field of medicine (Nesse and Williams, 1994), those inroads have been relatively recent. It is therefore likely that the concepts of (intralocus) sexual conflict and sexual antagonism are not generally well known within biomedical science, and variants that may have SA effects on disease risk may not be referred to as such. A secondary related hypothesis is that SA effects, when discovered, are discounted as errors as they do not match expectations that sex-differences in phenotype or genetic architecture are not important (Clayton and Collins, 2014). As a result, that may also lead them to be misclassified or simply go unreported, leading to a general publication bias.

For these reasons, we propose that a systematic review of the biomedical literature to identify SA loci in humans requires a specific and targeted set of search terms that would not normally be used within the field of evolutionary biology. We test this assumption by dividing our systematic review into two stages. In the first stage, we search for papers identifying specific genetic loci using terms directly relating to the concept of sexual antagonism and others utilized in evolutionary biology. We supplement this search with a second stage, where we develop a set of terms that we hypothesize may capture the same concept as sexual antagonism within the biomedical literature, although not explicitly stated as such, as well as additional terminology relating to possible alternatives for describing the two sexes. From our searches we also extracted data on sex-specific effect sizes and allele frequencies. We were then able to explore how effect sizes varied for the same alleles between the sexes, and test the prediction that SA alleles experiencing net positive selection between the sexes may achieve higher equilibrium frequencies than alleles that experience more symmetric or net negative patterns of selection (Morrow and Connallon, 2013). Our systematic review aims to advance our understanding of SA genes in humans, which show considerable and largely unexplained diversity in sexual dimorphism for disease phenotypes (Ober et al., 2006; Rigby and Kulathinal, 2015), but we also highlight how and why interdisciplinary research (in this case between biology and medicine) can sometimes fail, impeding scientific advances over significant periods of time.

## Methods

For this systematic review we followed PRISMA guidance where possible (Moher et al., 2009). PubMed (https://pubmed.ncbi.nlm.nih.gov/) was searched for articles on 2^nd^ December 2020 with no time limit. The searches were carried out in two Stages, with the organism filter set to human in both stages. In Stage 1, eligible studies were required to report specific genetic variants or haplotypes that were referred to as sexually antagonistic or were an example of intralocus sexual conflict. To achieve this, we conducted a Boolean search for articles that used the terms “sexual antagonism” OR “sexually antagonistic” OR “intralocus sexual conflict” AND “locus” OR “loci”, “gene” OR “snp” OR “polymorphism” OR “variant” OR “allele” in their abstract or title. The Stage 1 search returned 34 articles in total (full search term in the supplementary material; search output is accessible at https://pubmed.ncbi.nlm.nih.gov/collections/60255050/?sort=pubdate).

In Stage 2, studies were required to report specific genetic variants or haplotypes in humans with opposite effects in the two sexes on either complex traits, the outcome of a medical intervention, or disease risk/severity. We define complex traits as likely with a polygenic genetic architecture but are not directly related to a disease phenotype. In this second stage search terms were specifically designed to include papers from the biomedical literature that may have been missed in the first stage because they do not report their findings with terms normally found within the evolutionary biology literature. Again, we conducted a Boolean search for articles that used terms in their title or abstract to describe an opposite or different effect in the two sexes (“sex dependent”, “sex different”, “gender-dependent”, “sex AND opposite”, or “gender AND opposite”), or that capture this concept with alternative words for sex ((“male AND female AND opposite” OR “men AND women AND opposite” OR “boys AND girls AND opposite”) AND (“locus” OR “loci” OR “gene” OR “snp” OR “polymorphism” OR “variant” OR “allele”)). Full details of the search terms used and the numbers of articles returned are provided in the supplementary material. The Stage 2 search returned 881 papers (Figure 1) (full search term in the supplementary material; search output is accessible at https://pubmed.ncbi.nlm.nih.gov/collections/60254985/?sort=pubdate).

**Figure 1.**
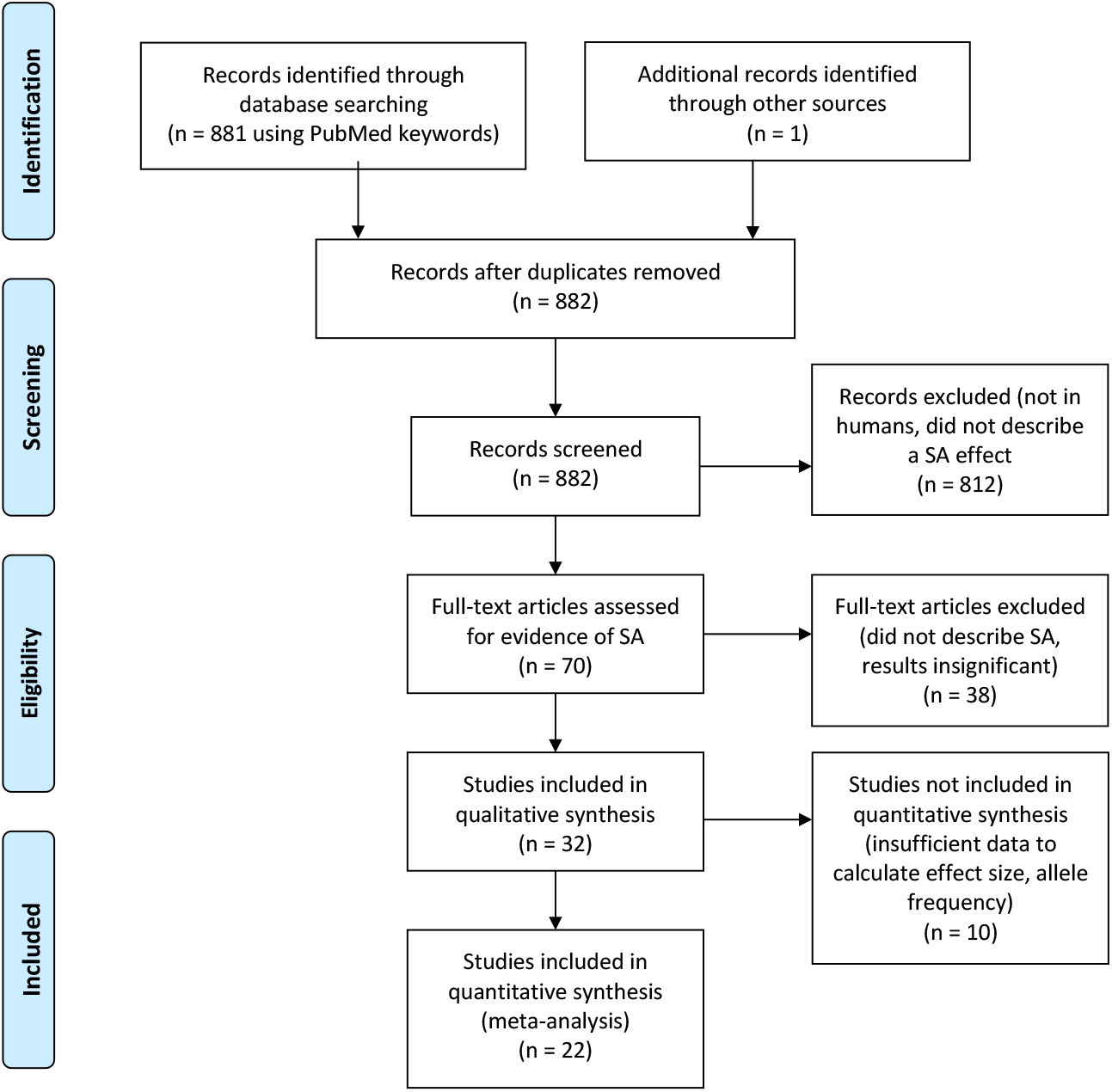
PRISMA flow diagram for systematic review of sexually antagonistic loci in humans.

The abstracts of the papers from Stage 1 were then examined by EM and from Stage 2 by JH, and any papers that had the possibility of reporting an opposite effect of a specific genetic locus on a complex trait, medical intervention or on disease risk/severity were considered for further screening. From Stage 1, no articles passed the screening. From Stage 2, this screening produced a shortlist of 70 candidate papers (https://pubmed.ncbi.nlm.nih.gov/collections/57906298/?sort=pubdate) Both JH and EM then reviewed all the full texts of these candidate papers in detail. Papers were included in the final list if they described a sex-opposite or SA effect linked to a specific genetic locus or loci, and reported the effect to be statistically significant (at a P-value cut-off of <0.05). One additional paper was considered from an outside source.

We converted all reported sex-specific effects into a standard effect size (Cohen’s *d*) quantifying the magnitude of how a given variant affects the studied trait expressed in the given sex. Specifically, Cohen’s *d* was computed based on the reported descriptive statistics (*N*, mean, standard error) or by conversion from other effect sizes (Odds ratio) and test statistics (*F*-values, *t*-values) using formulas reported elsewhere (Borenstein, 2009; Gurevitch et al., 2013, pp. 195–206; Lajeunesse, 2013). We sought information directly from the authors where these metrics were not possible to extract from the papers themselves (12 authors contacted, 5 responded, 3 responded with data, all later excluded as they did not fulfil the criteria for eligibility). We also recorded the gene name, locus (with accession/rs number where possible), trait affected and the frequency of the focal allele having the effect, hereafter referred to as *effect allele frequency*. Identification for the variants were taken from the studies where possible, but others necessitated searching the National Centre for Biotechnology Information (NCBI) to find the Reference SNP cluster ID for the variants described. Where effect allele frequencies were not reported, we used genotype frequencies to calculate effect allele frequency. Not all studies reported allele or genotype frequencies and so we attempted to supplement these data with allele frequencies from the 1000 genomes database (Auton et al., 2015), since these show a strong correlation with effect allele frequencies reported in the studies reviewed (Pearson correlation: *N* = 25 *r* = 0.94, *P* > 0.001; Figure S1). However, using this approach we were only able to supplement our allele frequency data for one additional locus (rs7341475 in the *RELN* gene; Table 1).

**Table 1.** Genetic loci in humans showing sexually antagonistic effects on trait expression. EAF, effect allele frequency (* indicates allele frequency derived from 100 genomes database); M, male; F, female. Background colours denote instances where genes and/or variants appear more than once in the list.

| Trait | Gene | Locus | EAF | M Effect | M Effect SE | F Effect | F Effect SE | Reference |
| --- | --- | --- | --- | --- | --- | --- | --- | --- |
| **Complex traits** |  |  |  |  |  |  |  |  |
| Concentration performance score | FADS1/2 | rs1535 | 0.672 | 0.2275 | 0.1065 | -0.2394 | 0.1116 | (Lauritzen et al., 2017) |
| Reading speed, number of sentences | FADS1/2 | rs174448 | 0.642 | 0.5511 | 0.1069 | -0.6074 | 0.1129 | (Lauritzen et al., 2017) |
| Reading speed, number of sentences | ELOVL5 | rs2397142 | 0.695 | -0.1963 | 0.1042 | 0.2364 | 0.1095 | (Lauritzen et al., 2017) |
| Reading speed, correct sentences | FADS1/2 | rs174448 | 0.642 | 0.6599 | 0.1077 | -0.3451 | 0.1112 | (Lauritzen et al., 2017) |
| Speed-dating success | OPRM1 | rs1799971 | 0.922 | 0.4697 | 0.2275 | -0.53 | 0.2856 | (Wu et al., 2016) |
| Speed-dating success | HTR2A | rs6311 | 0.418 | -0.4826 | 0.1982 | 0.4702 | 0.1964 | (Wu et al., 2016) |
| Waist-hip-ratio (BMI adjusted) | SIM1 | rs17185536 | 0.76 | -0.0166 | 0.008 | 0.0264 | 0.0067 | (Winkler et al., 2015) |
| Waist-hip-ratio (BMI adjusted) | NMU | rs3805389 | 0.28 | -0.0139 | 0.0067 | 0.0261 | 0.006 | (Winkler et al., 2015) |
| Waist-hip-ratio (BMI adjusted) | GNPNAT1 | rs4898764 | 0.53 | 0.0208 | 0.0066 | -0.0165 | 0.0059 | (Winkler et al., 2015) |
| Waist-hip-ratio (BMI adjusted) | SGCZ | rs17470444 | 0.71 | 0.0345 | 0.0081 | -0.0149 | 0.0068 | (Winkler et al., 2015) |
| Waist-hip-ratio (BMI adjusted) | IQGAP2 | rs2069664 | 0.53 | 0.0244 | 0.0078 | -0.0183 | 0.0067 | (Winkler et al., 2015) |
| Waist-hip-ratio (BMI adjusted) | SLC2A3 | rs741361 | 0.6 | 0.0272 | 0.0079 | -0.0196 | 0.0067 | (Winkler et al., 2015) |
| Waist-hip-ratio (BMI adjusted) | IRS1 | rs2673140 | 0.38 | 0.022 | 0.0066 | -0.0209 | 0.0059 | (Winkler et al., 2015) |
| Waist-hip-ratio (BMI adjusted) | TTN | rs2042995 | 0.23 | 0.0198 | 0.0066 | -0.022 | 0.0078 | (Winkler et al., 2015) |
| Waist-hip-ratio (BMI adjusted) | RXRA | rs10881574 | 0.07 | 0.0268 | 0.008 | -0.0191 | 0.0067 | (Winkler et al., 2015) |
| Waist-hip-ratio (BMI adjusted) | PTPRD | rs7042428 | 0.98 | 0.0244 | 0.0071 | -0.0166 | 0.0063 | (Winkler et al., 2015) |
| Waist-hip-ratio (BMI adjusted) | CECR2 | rs17809093 | 0.04 | 0.0294 | 0.0091 | -0.024 | 0.0077 | (Winkler et al., 2015) |
| Reaction time (RT) in the Stroop task, congruent condition | CHRNA5 | rs3841324 | 0.406 | -1.5421 | 0.3857 | 0.9945 | 0.2517 | (Schneider et al., 2015) |
| Reaction time (RT) in the Stroop task, incongruent condition | CHRNA5 | rs3841324 | 0.406 | -1.6215 | 0.389 | 1.0215 | 0.2521 | (Schneider et al., 2015) |
| Reaction time (RT) in the Negative priming task | CHRNA5 | rs3841324 | 0.406 | -1.675 | 0.3912 | 0.6299 | 0.247 | (Schneider et al., 2015) |
| Borderline personality traits after maltreatment | OXTR | rs53576 | 0.738 | -0.3429 | 0.0881 | 0.2066 | 0.0857 | (Cicchetti et al., 2014) |
| Borderline personality traits after maltreatment | FKBP5 | CATT haplotype | - | 0.2041 | 0.0877 | -0.195 | 0.0857 | (Cicchetti et al., 2014) |
| fluid intelligence | C16orf62 (OXTR) | rs226849 | 0.46 | -0.3949 | 0.1869 | 0.494 | 0.1814 | (Kiy et al., 2013) |
| face perception | C16orf62 (OXTR) | rs226849 | 0.46 | -0.5819 | 0.1909 | 0.3498 | 0.1788 | (Kiy et al., 2013) |
| face memory | C16orf62 (OXTR) | rs226849 | 0.46 | -0.3679 | 0.1864 | 0.3562 | 0.1789 | (Kiy et al., 2013) |
| Hippocampal volume | SLC6A4 | rs25531 | 0.39 | - |  | - |  | (Price et al., 2013) |
| Sociopathy (in alcoholics) | SLC6A4 | rs25531 | 0.51 | - |  | - |  | (Herman et al., 2011) |
| systolic blood pressure S_NIGHT | PON1 | rs3917542 | - | -0.1047 | 0.0637 | 0.1129 | 0.0645 | (Padmanabhan et al., 2010) |
| systolic blood pressure S_NIGHT | PON3 | rs17884563 | - | -0.1482 | 0.0637 | 0.1061 | 0.0645 | (Padmanabhan et al., 2010) |
| systolic blood pressure S_CLIN | EDNRB | rs1335803 | - | -0.1308 | 0.0637 | 0.1129 | 0.0645 | (Padmanabhan et al., 2010) |
| diastolic blood pressure D_NIGHT | DRD1 | rs265974 | - | -0.1482 | 0.0637 | 0.1502 | 0.0646 | (Padmanabhan et al., 2010) |
| diastolic blood pressure D_HOME | SLC4A2 | rs2303933 | - | -0.1482 | 0.0637 | 0.1325 | 0.0646 | (Padmanabhan et al., 2010) |
| diastolic blood pressure D_CLIN | LPL | rs17410577 | - | -0.1047 | 0.0637 | 0.1775 | 0.0647 | (Padmanabhan et al., 2010) |
| Psychological Stress | BDNF | rs6265 | 0.149 | - |  | - |  | (Shalev et al., 2009) |
| BMI standard deviation score (children) | FAS | rs2228305 | 0.0312 | -0.552 | 0.2259 | 0.4427 | 0.2075 | (Korner et al., 2007) |
| Pain sensitivity | OPRM1 | rs1799971 | 0.8817 | - |  | - |  | (Fillingim et al., 2005) |
| Central DBP | REN | rs6693954 | 0.22 | -0.4444 | 0.2132 | 1.1052 | 0.3923 | (Ljungberg et al., 2014) |
| **<u>Disease risk/severity</u>** |  |  |  |  |  |  |  |  |
| Graves ophthalmopathy in GD | PRR3 | rs2074503 | 0.3586 | 0.4397 | 0.1682 | -0.2185 | 0.0851 | (Liu et al., 2014) |
| Colorectal cancer | LTA | rs1041981 | 0.32 | -0.0722 | 0.0368 | 0.1027 | 0.0453 | (Sainz et al., 2012) |
| Arterial blood pressure | PPAR | rs1801282 | 0.1213 | - |  | - |  | (Franck et al., 2012) |
| Schizophrenia risk | RELN | rs7341475 | 0.851* | 1.2396 | 0.2819 | -0.762 | 0.3662 | (Ovadia and Shifman, 2011) |
| Colon cancer survival | ESR2 | (CA)n repeats | - | 0.2272 | 0.0962 | -0.2046 | 0.1205 | (Press et al., 2011) |
| Schizophrenia risk | DAO | rs2070586 | 0.2729 | 0.1982 | 0.0888 | -0.2537 | 0.0908 | (Kim et al., 2010) |
| T2DM, fasting glucose | SCARB1 | rs9919713 | 0.04 | 0.1865 | 0.0853 | -0.2515 | 0.0697 | (McCarthy et al., 2009) |
| Depression | SLC6A4 | rs25531 | 0.4586 | 0.5404 | 0.2277 | -0.3996 | 0.1859 | (Sjöberg et al., 2006) |
| Infertility in cystic fibrosis | CFTR | rs213950 | 0.417931 | - |  | - |  | (Morea et al., 2005) |
| Bone mineral density | APOE | rs429358 | 0.0798 | - |  | - |  | (Wong et al., 2005) |
| Narcolepsy severity | COMT | Long allele | 0.4588 | 1.3596 | 0.3878 | -2.2152 | 0.4871 | (Dauvilliers et al., 2001) |
| Nonsyndromic cleft lip | IMPK | rs11142081 | 0.863 | 1.2156 | 0.0937 | -1.1161 | 0.0783 | (Carlson et al., 2018) |
| Nonsyndromic cleft lip | IMPK | rs72804706 | 0.898 | -0.816 | 0.0906 | 2.0665 | 0.0908 | (Carlson et al., 2018) |
| Nonsyndromic cleft lip | IMPK | rs77590619 | 0.744 | -0.8577 | 0.0904 | 1.2291 | 0.0804 | (Carlson et al., 2018) |
| Allergy | ACP1 | A* allele (CG haplotype) | - | - |  | - |  | (Gloria-Bottini et al., 2007) |
| Hepatitis C susceptibility | IL-12B | rs3212227 | 0.3463 | - |  | - |  | (Youssef et al., 2013) |
| RSV severity | IL-9 | rs2069885 | 0.15 | 0.1888 | 0.0933 | -0.2794 | 0.0957 | (Schuurhof et al., 2010) |
| CV risk marker (plasma fatty acid composition) | FABP2 | rs1799883 | 0.287 | - |  | - |  | (Gastaldi et al., 2007) |
| Bone mineral density | NR3C1 | GCAG haplotype | - | 0.6854 | 0.3676 | -0.9029 | 0.4311 | (Peng et al., 2008) |
| Anxiety | NGF | rs6330 | 0.451 | 0.4562 | 0.18 | -0.4674 | 0.1817 | (Lang et al., 2008) |
| Melanoma risk | RNASEL, miR-146a | AG/AA and C haplotype | - | 0.4525 | 0.1564 | -0.3553 | 0.174 | (Sangalli et al., 2017) |

If *b*_f_ and *b*_m_ represent the estimated effect of the focal allele in females and males respectively, then positive values of *b*_f_ or *b*_m_ imply that the allele is positively associated with disease expression, whereas negative values imply that the allele is negatively associated with disease expression. SA alleles are therefore defined as those with opposite effects between the sexes (i.e. *b*_f_ < 0 < *b*_m_ or *b*_m_ < 0 < *b*_f_). An evolutionary model of evolution at SA loci predicts that as the magnitude of the positive effect that an SA allele has outweighs the negative effect, the higher the frequency that allele can achieve (Morrow and Connallon, 2013). We therefore expect a negative relationship between the ratio of positive to negative effect sizes (hereafter referred to as the *effect size ratio*) and observed allele frequency. To investigate this, we calculated the effect size ratio by dividing the positive standardized effect size by the negative standardized effect size; when *b*_f_ < 0 < *b*_m_ effect size ratio was calculated as *b*_m_/ *b*_f_, and when *b*_m_ < 0 < *b*_f_ effect size ratio was calculated as *b*_f_/ *b*_m_.

Thus, SA alleles with a greater beneficial effect will have a smaller, more negative effect size ratio (<-1), while SA alleles that have a greater deleterious effect will have a larger effect size ratio (>-1), up to a maximum value of 0 (positive values occur when the effect is positive or negative in both sexes, but the allele would then no longer be defined as a SA allele).

We modelled how effect allele frequency changes with effect size ratio using a generalized linear model (GLM), weighted by the inverse of the variance of the effect sizes such that data points with smaller variance have a higher weight, since smaller sample sizes were associated with larger and more variable effect sizes (Figure S2). We initially also included trait class and its interaction with effect size ratio as a fixed factor with two levels (complex trait and disease trait/severity), since alleles that influence complex traits in opposite directions are not necessarily under SA selection and so may not behave in the way predicted, whereas alleles influencing disease traits, unless very late acting, are more likely to show a closer relationship with marginal effects on fitness. As a response variable, effect allele frequency is limited between 0 and 1, so we looked at allele counts to allow allele frequency to vary freely. GLMs with binomial error distribution showed substantial overdispersion, so we used a quasibinomial function to address this issue (Payne et al., 2018). We used a Chi-squared test to infer significance of the two predictor variables and their interaction. The full model indicated that the interaction term and trait class have no significant effect, so that we report the fit of the reduced model which only includes effect size ratio. We also subsequently modelled the data for the two trait classes (complex traits and disease risk/severity) separately, again using a quasibinomial distribution function, to see if the result was replicated in these smaller subsets of the data. The raw data and R script is available for replicating the analyses and figures we present (Supplementary Material).

## Results

The Stage 1 search found no articles that described genetic loci in humans with effects that were sexually antagonistic. In contrast, the Stage 2 search identified 32 articles that described variants with statistically significant sex-opposite or SA effects (Figure 1; https://pubmed.ncbi.nlm.nih.gov/collections/60278165/?sort=pubdate). From the studies examined, 51 SA variants were identified (Table 1), affecting 21 different complex traits (30 loci) and 19 disease risk/severity traits (21 loci), with a large range of effect sizes. There were no variants affecting medical intervention traits that passed the screening process. The majority of alleles had effect sizes of similar absolute values in the two sexes, with larger effects tending to show greater differences on absolute effect size (Figure S3).

We found 22 studies provided sufficient data to allow the relationship between effect size ratio and effect allele frequency to be explored by statistical modelling. Trait class and its interaction with effect size ratio were initially included in the model but neither were found to have a significant effect (GLM: trait class dfs = 1,35, deviance = 15.5 × 10^3^, *P* = 0.965; interaction dfs = 1,34, deviance = 1476, P = 0.989) and were therefore excluded from the subsequent model in which effect size ratio was found to relate negatively to effect allele frequency, as predicted (GLM: estimate ± SE = −1.10 ± 0.50, dfs = 1,36, deviance = 38.7 × 10^6^, *P* = 0.024; Figure 2). For three loci there were data on effect sizes for more than one trait (Table 1), which introduced a degree of non-independence between these values in the dataset. However, resampling the data with effect sizes for only one trait for these three loci in turn and modelling these datasets in the same way (18 individual GLMs, dfs = 1,31) did not change the results qualitatively (minimum, maximum: model estimate = −1.0963, −1.0964; deviance = 38.72 × 10^6^, 38.73 × 10^6^; *P* = 0.03618, 0.03620).

**Figure 2.**
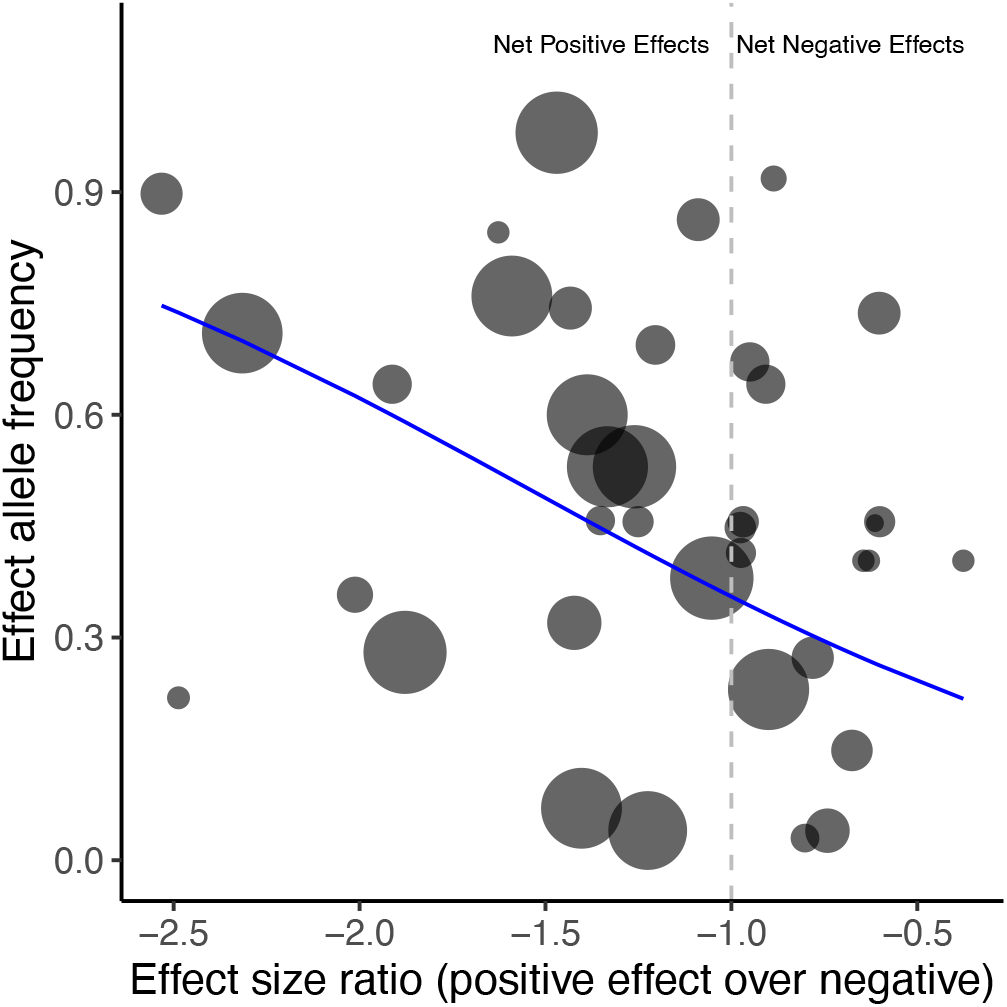
The relationship between effect allele frequency and effect size ratio. Point size varies according to the variance of effect size ratio - larger points have smaller variance and therefore a larger weighting in the model. The vertical dotted line represents the switch point between a net-negative effect of a particular locus (effect size ratio >-1) and a net positive effect (effect size ratio < −1). The blue line represents the predicted values derived from the generalized linear model fitted to the data (see Results).

Although the interaction term between trait type and effect size ratio was not significant we wanted to investigate whether the negative relationship between effect size ratio and effect allele frequency across all loci was repeated when the data was divided according to trait class, since while opposite effects on disease risk/severity may show a direct relationship with SA fitness effects, complex traits may or may not be experiencing SA selection, even if effects are in opposite directions in the two sexes. However, although the trends were both again negative as predicted, the generalized linear models of these smaller datasets were only marginally significant (complex traits model estimate ± SE = −1.10 ± 0.61, dfs = 1,24, deviance = 38.7 × 106, P = 0.065; disease risk/severity model estimate ± SE = −1.34 ± 0.77, dfs = 1,10, deviance = 5.14 × 104, P = 0.053; Figure S4). As for the full dataset, there was some lack of independence for the complex traits subset which contained loci with effect sizes for more than one trait. Again, resampling and modelling these datasets in the same way (18 individual glms, dfs 1, 22) did not change the results quantitatively (minimum, maximum: model estimate = −1.0960, −1.0961; deviance = 38.669 × 10^6^, 38.675 × 10^6^; *P* = 0.07774, 0.07776).

## Discussion

The Stage 1 literature search of our systematic review focussed on finding reports of specific genetic loci in humans with evidence for SA effects using terminology normally associated with the evolutionary concepts of sexual conflict or sexual antagonism. The Stage 2 search sought equivalent evidence, but used search terms that we anticipated would be used by scientists outside the field of evolutionary biology, who may not use the same terminology. Although the Stage 1 search did not find any examples of SA loci occurring in humans, the Stage 2 search identified 51 genetic loci across 39 studies that had ostensibly SA effects, but were not described as such. Clearly then the Stage 1 review failed to identify multiple relevant reports going back some 20 years because the terminology used in those reports did not match the conceptual framework of the search. The Stage 2 search may also be a lower limit given searches of other larger databases may also harbour further examples (see below). Although not yet validated, these reports nonetheless represent a substantial body of evidence that humans, like other organisms with separate sexes (Bonduriansky and Chenoweth, 2009), inevitably experience SA selection for a wide range of complex traits (Connallon and Clark, 2014; Garver-Apgar et al., 2011; Stearns et al., 2012; Stulp et al., 2012), as well as for a range of diseases.

A key prediction from a population genetic model of SA genetic variation is that SA alleles are expected to achieve higher equilibrium frequencies, without necessarily going to fixation, as the relative magnitude of the positive effect in one sex outweighs the negative effect in the opposite sex i.e. an increasingly negative effect size ratio (Morrow and Connallon, 2013). We found clear support for this prediction, with a negative relationship between effect size ratio and allele frequency when examining all complex traits and diseases together (Fig 2), although this relationship was only marginally significant when complex or disease trait classes were modelled separately (Fig S3). These results lend support to the view that the loci identified are genuinely experiencing SA selection, and although it is difficult to discern how selection acts on trait size in males and females for complex traits we would not expect the frequencies of disease-causing alleles to be so high under a mutation-selection balance. We were not able to look at the evolutionary history or age of these alleles due to limited information (n = 8) (Albers and McVean, 2018), it would nonetheless be valuable to examine the population dynamics of these alleles over a broad timescale.

The loci showed a very broad range of effect sizes, from small to very large, with a generally symmetrical inverse relationship between effect size in one sex and the other (Figure S2), which is generally expected, since our screening process necessarily excluded studies reporting effects in the same direction across the two sexes. Nonetheless, it is striking just how large some of the effects were, with very large negative effects in one sex simultaneously occurring with similarly large positive effects. Larger and more variable effect sizes are expected when sample sizes are small, a pattern we also found, which motivates further investigation of the traits and loci included in this review to elucidate more accurate estimates of real effect sizes.

The loci themselves fall within genes that influence a broad range of phenotypes, including morphological, physiological and behavioural complex traits, and a similarly diverse range of disease types, including various cancers, neurological disorders and immune system processes (see Table 1). For the most part, the diseases appear to be early-acting rather than late-onset, with the exception of perhaps bone mineral density. As such, it seems reasonable that an allele that increases disease risk or severity in these cases will indeed have a concomitant reduction in marginal fitness. Although a single large study identified several loci related to BMI adjusted waist-to-hip ratio, there is generally no overall bias towards one particular trait or disease class. The total number of loci is relatively small however, which may limit power to identify such biases if they exist. We also did not find any examples of SA loci on the X-chromosome, which may or may not be a hotspot of sexual antagonism (Fry, 2010; Rice, 1984; Ruzicka and Connallon, 2020). This was expected given that it is commonly not included in genome-wide analyses (Wise et al., 2013), although most of the papers we reviewed were candidate gene studies. Nonetheless, we anticipate further examples may be identified should data from the X-chromosome be included systematically in association studies.

We also found evidence that some traits are influenced by more than one genetic variant in sex-opposite or SA ways (n=8), or that some specific variants have pleiotropic effects on more than one trait or disease in a sex-opposite or SA way (n=6; see Table 1). This complexity in the genetic architecture of traits or diseases may make conflict resolution particularly difficult, as a change in the allele at a single genetic locus experiencing SA selection may simultaneously influence multiple genes or phenotypes in both sexes in divergent ways (Fitzpatrick, 2004; Pennell and Morrow, 2013). Consequently, this may explain the persistence of intralocus sexual conflict at these loci. It may be that some of the remaining loci also have as yet unidentified pleiotropic effects with other traits or there are as yet unidentified SA loci influencing those same traits or diseases. Of the 51 variants identified, 4 were confirmed to be haplotypes. Such variants, consisting of two or more SNPs in linkage disequilibrium having a joint effect, could also present a problem for conflict resolution, since the ability of selection to act on any individual locus independent of the others in the linkage block is reduced for linked loci. This issue may also extend to other single variants reported here if they also occur in linkage blocks.

Although we have identified multiple genetic loci with either sex-opposite or SA effects these are likely outnumbered by those with either sex-specific (same direction but different magnitude) or sex-limited effects. For instance, Winkler et al. (2015) report 44 loci with sex-specific or sex-limited effects but only 11 with sex-opposite effects. Several other genome-wide association studies report sex-specific effects in several human diseases (Khramtsova et al., 2019), and a recent (non-systematic) review identified 37 SNPs with sex-dependent effects (Gilks et al., 2014). Detection of loci with SA versus sex-specific or sex-limited effects may differ systematically since we expect SA loci are more likely to persist at intermediate frequencies than loci with sex-specific effects (i.e. that differ between the sexes in magnitude but not sign). A larger systematic review that targeted variants with sex-specific or sex-dependent effects would therefore be a valuable contribution and enable us to more clearly understand how important SA alleles are relative to the broader context of genes with non-identical effects in the two sexes.

A key gap in our knowledge is whether the putative examples of sex-opposite or SA alleles presented here can be validated using independently derived datasets in the same or different sub-populations. For this, focussing on those variants associated with sex-differential disease risk would naturally be the most fruitful for advancing our knowledge of sexual antagonism in human disease. We therefore encourage specialists for the particular disease groups presented in Table 1 to include in the future the potential for SA genetic effects to occur when designing studies and analysing data. It is also important for all studies to report results in sufficient detail so that effect sizes can be calculated (if not given) as well as allele frequency data. Unfortunately, 17 studies failed to do this, so could not be included in our review and subsequent test of the evolutionary model. More generally, our study reinforces the view that sex is an important factor in shaping genetic associations with human traits and disease, even in divergent and contradictory ways, and so should always be considered when investigators examine genetic associations with phenotypes (Lee, 2018; Ober et al., 2008).

There is a possibility that some bias exists in the PubMed database against papers from the field of evolutionary biology, and if we had used another database then both stages would have returned valid papers. We subsequently (14^th^ Dec 2020) sought to investigate this possibility with additional searches of Scopus and Web of Science. Repeating the Stage 1 search initially returned 85 papers in Web of Science (Basic search of Topic, refined by “Human” within results) and 71 papers in Scopus (include keywords “Human”, exclude “Non-Human”), but again no papers made it through the screening process. Repeating the Stage 2 search returned a much larger number of studies in both Web of Science (4,573; Basic search of Topic, refined by “Human” within results) and Scopus (6,616; Advanced search of Title-Abstract, filter: include human and humans, exclude nonhumans, Articles only), with 8,604 unique records. This large number of records is too many to currently screen and may harbour further examples of SA loci in humans, but it is noteworthy that the majority of the initial list of studies found during Stage 2 using PubMed were also found in the other two databases (712 out of 881) as well as 28 out of the screened list of 32 studies identified in the current systematic review.

The disparity between the two stages of the review process provides a remarkable example of how discipline-specific terminology and concepts can hinder scientific communication between fields for substantial periods of time. For many of the studies identified in this review, the focus of the research was not on quantifying or even identifying sex-specific genetic effects in traits or disease, although they were tested for. The responses to such findings varied. Some framed these results as a major finding, others merely made note of them, with one suggesting that since the genetic associations were in opposite directions in the two sexes then it should be regarded as a false positive (Wong et al., 2005). It is possible, or even likely then that there exists a publication bias in the biomedical sciences against studies with apparently incongruous sex-opposite or SA effects, perhaps in part because the evolutionary framework that may provide a context for understanding those results is not generally appreciated or known. Of course, for evolutionary biologists interested in the role of SA selection on human traits and disease, this is clearly a problem. For many years no specific examples of SA loci were known in humans, and yet the first putative examples in fact appeared in the literature two decades ago. Our systematic review therefore provides a concrete illustration of how advances to both fields have likely been hampered by a limited understanding of the theoretical framework and terminology used by the neighbouring field.

How can communication between the fields of medicine and evolutionary biology be facilitated? We do not have a comprehensive answer but clearly training in, or an increased awareness of, evolutionary medicine for scientists in both camps would probably be beneficial. The topic of evolutionary biology now forms a key part of the education for some biomedical scientists and clinicians. Similarly, evolutionary medicine is now frequently included in fundamental textbooks on evolution, and is being taught to students of evolutionary biology as part of their degree courses. These changes have been relatively recent, are far from being universally employed, but they should be encouraged. Funders of primary research are also now actively encouraging or obliging researchers to include sex as a factor when designing their studies (Clayton and Collins, 2014), although this may simply result in studies that aim to control for rather than investigate these effects explicitly. Consequently even this remedy may still preclude a deeper understanding of the origins of some human diseases.

## Supporting information

Raw data

## Data Availability

All the data is publicly available on the Pubmed database. Links to specific collections of articles are all given in the manuscript. These are:
https://pubmed.ncbi.nlm.nih.gov/collections/60255050/?sort=pubdate
https://pubmed.ncbi.nlm.nih.gov/collections/60254985/?sort=pubdate
https://pubmed.ncbi.nlm.nih.gov/collections/57906298/?sort=pubdate
https://pubmed.ncbi.nlm.nih.gov/collections/60278165/?sort=pubdate
Data for the meta-analysis is provided as a file in the supplementary material.

## Acknowledgements

We thank Jessica Abbott, Tim Connallon and Filip Ruzicka for comments on a previous version of the MS. Funding was provided by the Swedish Research Council (Grant number: 2019-03567) and by the Royal Society to EHM as a University Research Fellowship and Enhancement Award. TJ was funded by the Centre national de la recherche scientifique (CNRS) and the German Research Foundation (DFG grant number: JA 2653/2-1).

## Supplementary Material

Both stages of the PubMed literature search employed a Species filter set to Humans only.

### Stage 1 Search term

((((((((((((((((((((sexual antagonism[Title/Abstract] AND locus[Title/Abstract]) OR (sexual antagonism[Title/Abstract] AND loci[Title/Abstract])) OR (sexual antagonism[Title/Abstract] AND gene[Title/Abstract])) OR (sexual antagonism[Title/Abstract] AND snp[Title/Abstract])) OR (sexual antagonism[Title/Abstract] AND polymorphism[Title/Abstract])) OR (sexual antagonism[Title/Abstract] AND variant[Title/Abstract])) OR (sexual antagonism[Title/Abstract] AND allele[Title/Abstract])) OR (sexually antagonistic[Title/Abstract] AND locus[Title/Abstract])) OR (sexually antagonistic[Title/Abstract] AND loci[Title/Abstract])) OR (sexually antagonistic[Title/Abstract] AND gene[Title/Abstract])) OR (sexually antagonistic[Title/Abstract] AND snp[Title/Abstract])) OR (sexually antagonistic[Title/Abstract] AND polymorphism[Title/Abstract])) OR (sexually antagonistic[Title/Abstract] AND variant[Title/Abstract])) OR (sexually antagonistic[Title/Abstract] AND allele[Title/Abstract])) OR (intralocus sexual conflict[Title/Abstract] AND locus[Title/Abstract])) OR (intralocus sexual conflict[Title/Abstract] AND loci[Title/Abstract])) OR (intralocus sexual conflict[Title/Abstract] AND gene[Title/Abstract])) OR (intralocus sexual conflict[Title/Abstract] AND snp[Title/Abstract])) OR (intralocus sexual conflict[Title/Abstract] AND polymorphism[Title/Abstract])) OR (intralocus sexual conflict[Title/Abstract] AND variant[Title/Abstract])) OR (intralocus sexual conflict[Title/Abstract] AND allele[Title/Abstract])

### Stage 2 Search term

(((((((((((((((((((((((((((((((((((((((((((((((((((((((gender[Title/Abstract] AND opposite[Title/Abstract] AND locus[Title/Abstract]) OR (gender[Title/Abstract] AND opposite[Title/Abstract] AND loci[Title/Abstract])) OR (gender[Title/Abstract] AND opposite[Title/Abstract] AND gene[Title/Abstract])) OR (gender[Title/Abstract] AND opposite[Title/Abstract] AND snp[Title/Abstract])) OR (gender[Title/Abstract] AND opposite[Title/Abstract] AND polymorphism[Title/Abstract])) OR (gender[Title/Abstract] AND opposite[Title/Abstract] AND variant[Title/Abstract])) OR (sex[Title/Abstract] AND opposite[Title/Abstract] AND locus[Title/Abstract])) OR (sex[Title/Abstract] AND opposite[Title/Abstract] AND loci[Title/Abstract])) OR (sex[Title/Abstract] AND opposite[Title/Abstract] AND gene[Title/Abstract])) OR (sex[Title/Abstract] AND opposite[Title/Abstract] AND snp[Title/Abstract])) OR (sex[Title/Abstract] AND opposite[Title/Abstract] AND polymorphism[Title/Abstract])) OR (sex[Title/Abstract] AND opposite[Title/Abstract] AND variant[Title/Abstract])) OR (sex dependent[Title/Abstract] AND locus[Title/Abstract])) OR (sex dependent[Title/Abstract] AND loci[Title/Abstract])) OR (sex dependent[Title/Abstract] AND gene[Title/Abstract])) OR (sex dependent[Title/Abstract] AND snp[Title/Abstract])) OR (sex dependent[Title/Abstract] AND polymorphism[Title/Abstract])) OR (sex dependent[Title/Abstract] AND variant[Title/Abstract])) OR (sex different[Title/Abstract] AND locus[Title/Abstract])) OR (sex different[Title/Abstract] AND loci[Title/Abstract])) OR (sex different[Title/Abstract] AND gene[Title/Abstract])) OR (sex different[Title/Abstract] AND snp[Title/Abstract])) OR (sex different[Title/Abstract] AND polymorphism[Title/Abstract])) OR (sex different[Title/Abstract] AND variant[Title/Abstract])) OR (gender-dependent[Title/Abstract] AND locus[Title/Abstract])) OR (gender-dependent[Title/Abstract] AND loci[Title/Abstract])) OR (gender-dependent[Title/Abstract] AND gene[Title/Abstract])) OR (gender-dependent[Title/Abstract] AND snp[Title/Abstract])) OR (gender-dependent[Title/Abstract] AND polymorphism[Title/Abstract])) OR (gender-dependent[Title/Abstract] AND variant[Title/Abstract])) OR (male[Title/Abstract] AND female[Title/Abstract] AND opposite[Title/Abstract] AND locus[Title/Abstract])) OR (male[Title/Abstract] AND female[Title/Abstract] AND opposite[Title/Abstract] AND loci[Title/Abstract])) OR (male[Title/Abstract] AND female[Title/Abstract] AND opposite[Title/Abstract] AND gene[Title/Abstract])) OR (male[Title/Abstract] AND female[Title/Abstract] AND opposite[Title/Abstract] AND snp[Title/Abstract])) OR (male[Title/Abstract] AND female[Title/Abstract] AND opposite[Title/Abstract] AND polymorphism[Title/Abstract])) OR (male[Title/Abstract] AND female[Title/Abstract] AND opposite[Title/Abstract] AND variant[Title/Abstract])) OR (men[Title/Abstract] AND women[Title/Abstract] AND opposite[Title/Abstract] AND locus[Title/Abstract])) OR (men[Title/Abstract] AND women[Title/Abstract] AND opposite[Title/Abstract] AND loci[Title/Abstract])) OR (men[Title/Abstract] AND women[Title/Abstract] AND opposite[Title/Abstract] AND gene[Title/Abstract])) OR (men[Title/Abstract] AND women[Title/Abstract] AND opposite[Title/Abstract] AND snp[Title/Abstract])) OR (men[Title/Abstract] AND women[Title/Abstract] AND opposite[Title/Abstract] AND polymorphism[Title/Abstract])) OR (men[Title/Abstract] AND women[Title/Abstract] AND opposite[Title/Abstract] AND variant[Title/Abstract])) OR (boys[Title/Abstract] AND girls[Title/Abstract] AND opposite[Title/Abstract] AND locus[Title/Abstract])) OR (boys[Title/Abstract] AND girls[Title/Abstract] AND opposite[Title/Abstract] AND loci[Title/Abstract])) OR (boys[Title/Abstract] AND girls[Title/Abstract] AND opposite[Title/Abstract] AND gene[Title/Abstract])) OR (boys[Title/Abstract] AND girls[Title/Abstract] AND opposite[Title/Abstract] AND snp[Title/Abstract])) OR (boys[Title/Abstract] AND girls[Title/Abstract] AND opposite[Title/Abstract] AND polymorphism[Title/Abstract])) OR (boys[Title/Abstract] AND girls[Title/Abstract] AND opposite[Title/Abstract] AND variant[Title/Abstract])) OR (gender[Title/Abstract] AND opposite[Title/Abstract] AND allele[Title/Abstract])) OR (sex[Title/Abstract] AND opposite[Title/Abstract] AND allele[Title/Abstract])) OR (sex dependent[Title/Abstract] AND allele[Title/Abstract])) OR (sex different[Title/Abstract] AND allele[Title/Abstract])) OR (gender-dependent[Title/Abstract] AND allele[Title/Abstract])) OR (male[Title/Abstract] AND female[Title/Abstract] AND opposite[Title/Abstract] AND allele[Title/Abstract])) OR (men[Title/Abstract] AND women[Title/Abstract] AND opposite[Title/Abstract] AND allele[Title/Abstract])) OR (boys[Title/Abstract] AND girls[Title/Abstract] AND opposite[Title/Abstract] AND allele[Title/Abstract])

**Figure S1.**
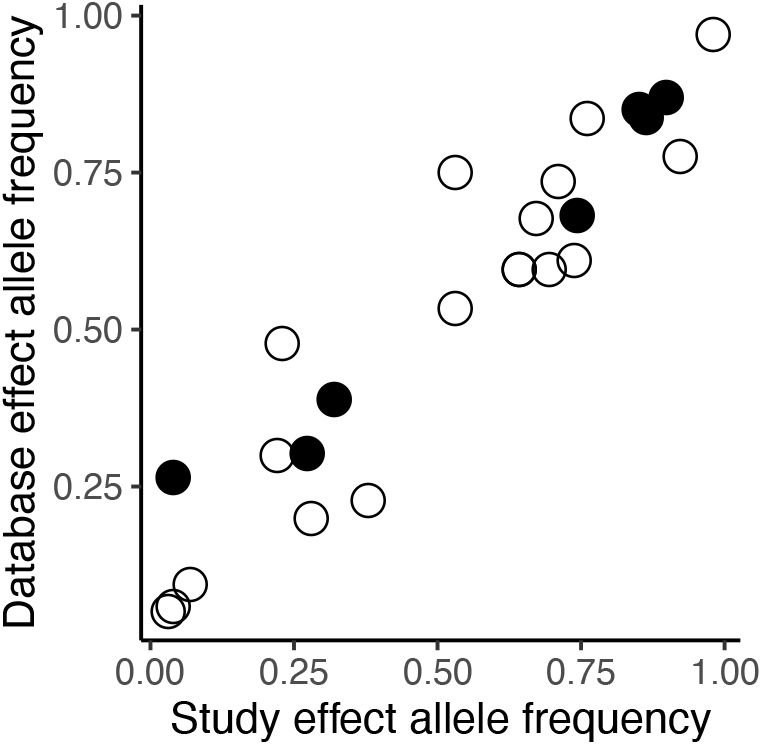
Study reported effect allele frequencies and database allele frequencies are strongly correlated. The correlation between reported effect allele frequency and frequencies for the same alleles obtained from the 1000 genomes database (0.94). Filled circles represent disease risk/severity related variants while open circles represent complex trait variants.

**Figure S2.**
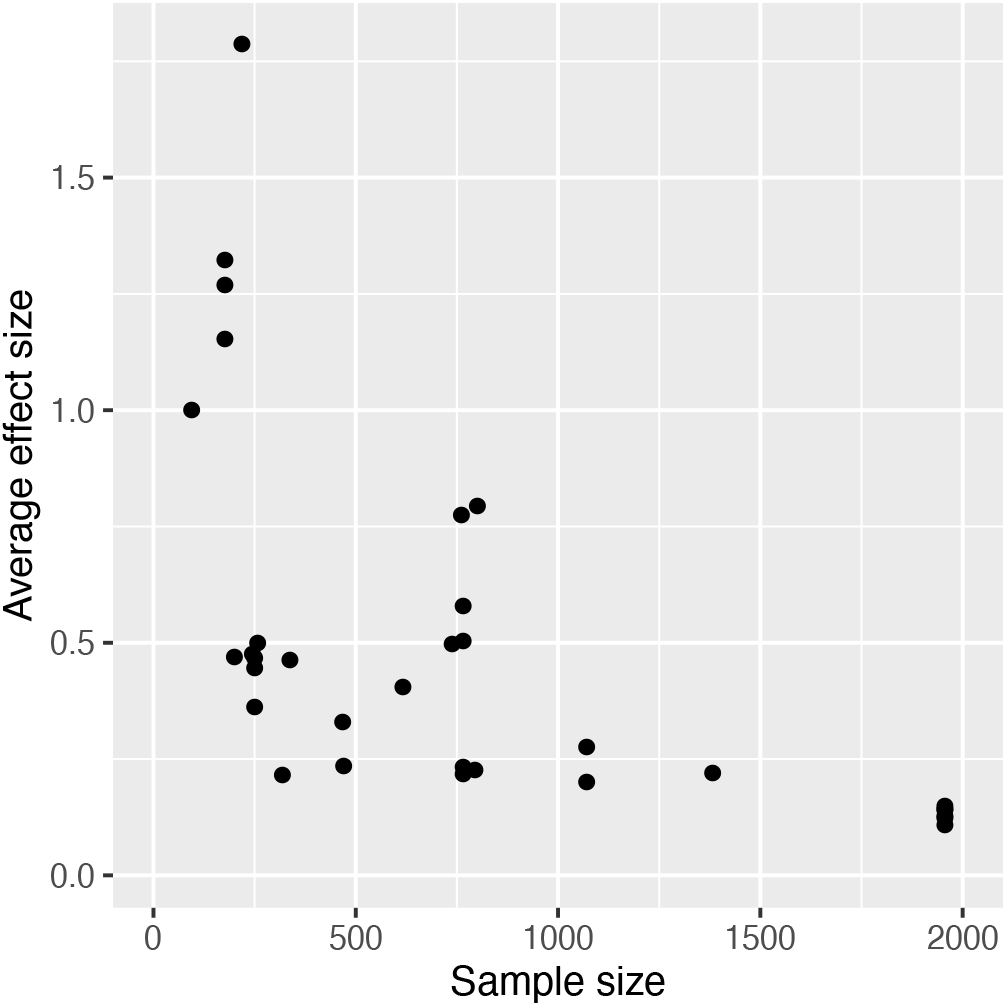
Relationship between study sample size and average extracted effect size across all studies.

**Figure S3.**
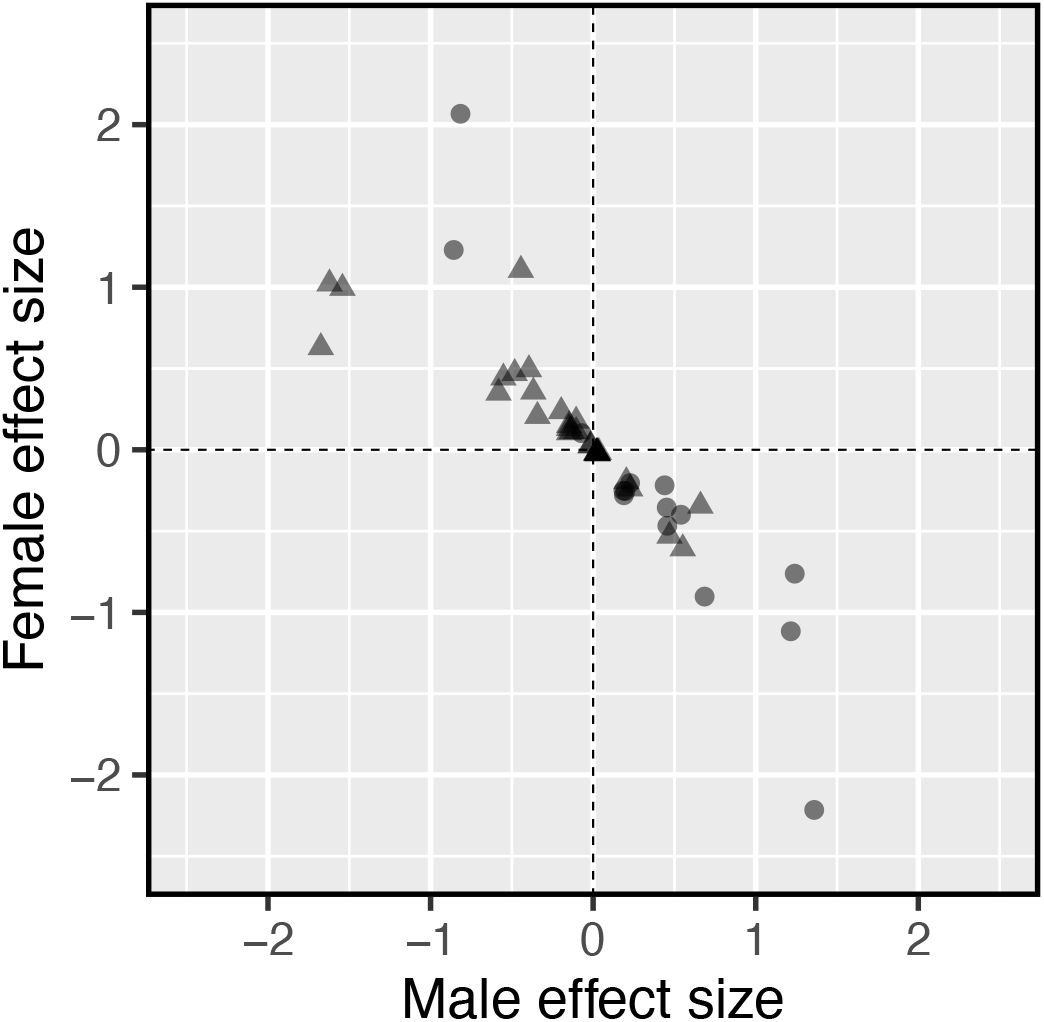
Female effect size and male effect size are negatively correlated in SA alleles. Effect size (Cohen’s D) of variants in females against effect size in males. Triangular points represent sex-opposite complex trait variants, circular points represent disease risk/severity variants.

**Figure S4.**
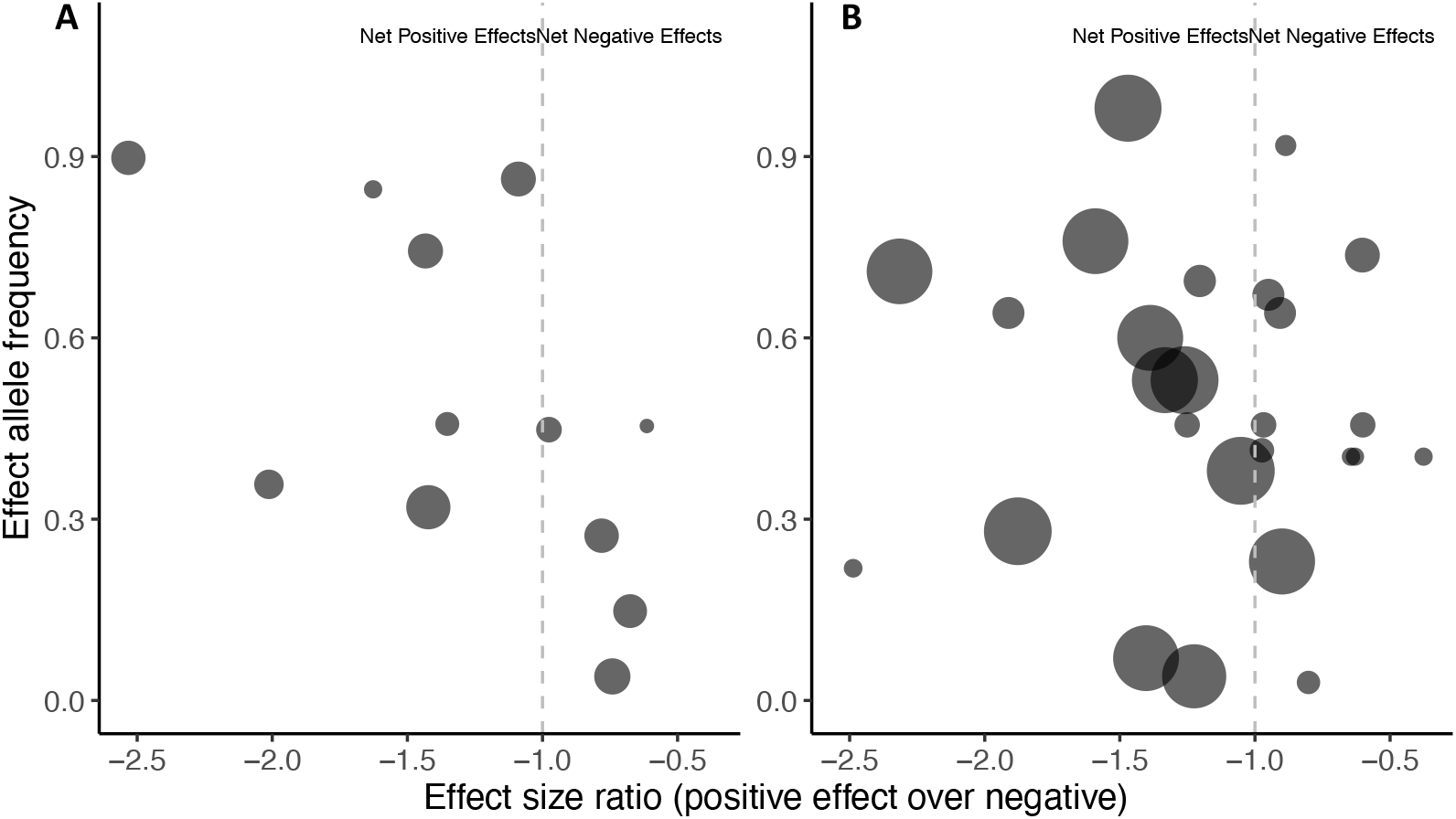
The relationship between effect allele frequency and effect size ratio, grouped by trait class. Points to the right of the vertical dotted line have a greater negative effect than positive. **A**. Disease risk/severity variants. **B**. Complex traits. Point size is based on the variance of the effect size ratio, with smaller variance having larger point sizes and greater weighting in the statistical model.

