## Supplementary material for "Systematic review reveals multiple sexually antagonistic polymorphisms affecting human disease and complex traits"

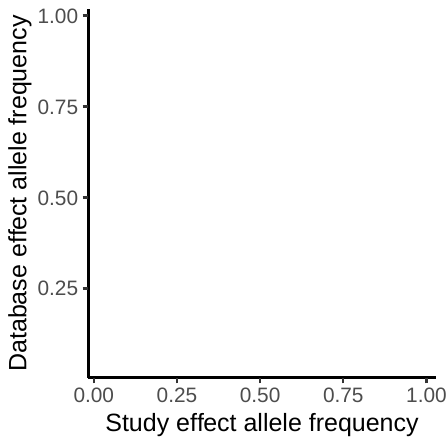

**Figure S1. Study reported effect allele frequencies and database allele frequencies are strongly correlated.** The correlation between reported effect allele frequency and frequencies for the same alleles obtained from the 1000 genomes database (0.94). Filled circles represent disease risk/severity related variants while open circles represent complex trait variants.

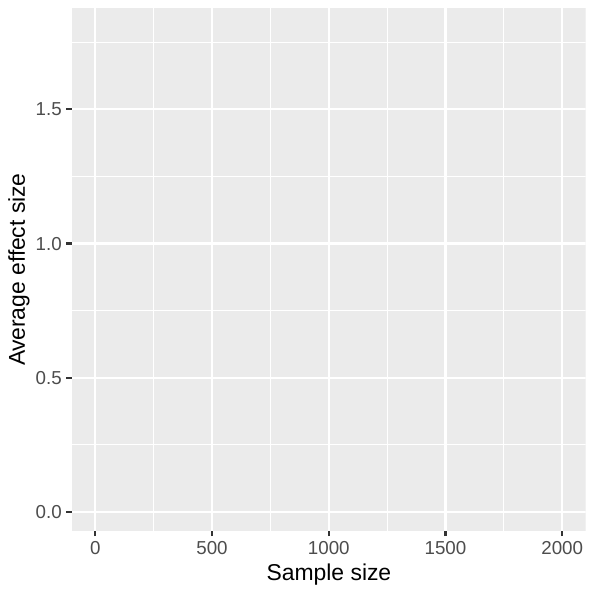

**Figure S2. Relationship between study sample size and average extracted effect size across all studies.**

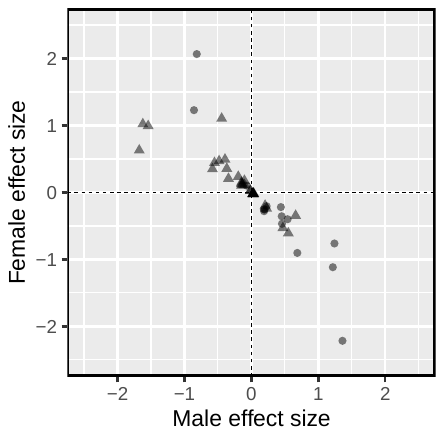

**Figure S3. Female effect size and male effect size are negatively correlated in SA alleles.** Effect size (Cohen’s D) of variants in females against effect size in males. Triangular points represent sex-opposite complex trait variants, circular points represent disease risk/severity variants.

**B**

**A**

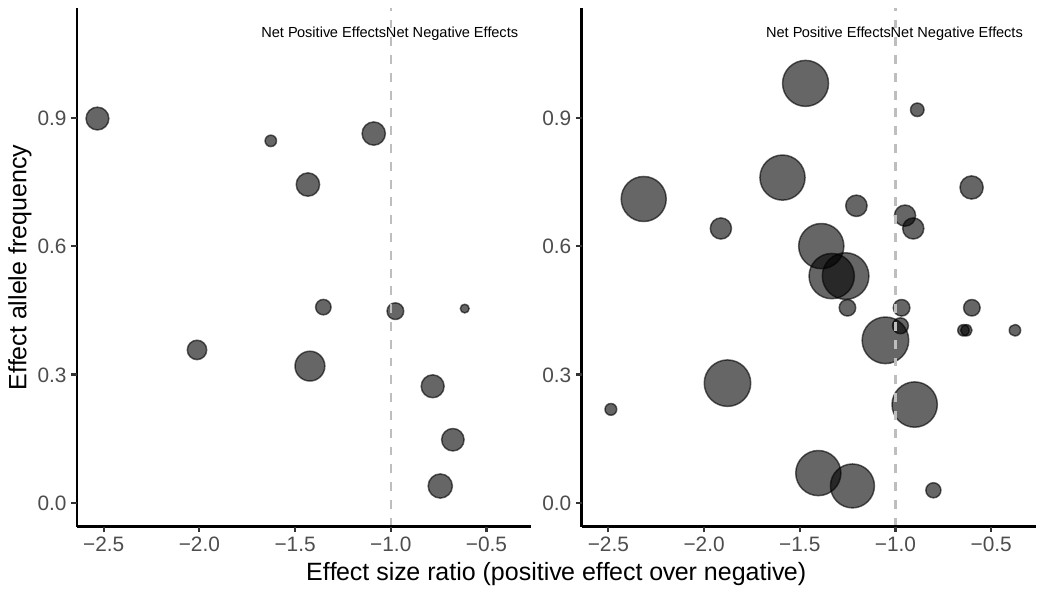

**Figure S4. The relationship between effect allele frequency and effect size ratio, grouped by trait class.** Points to the right of the vertical dotted line have a greater negative effect than positive. **A.** Disease risk/severity variants. **B.** Complex traits. Point size is based on the variance of the effect size ratio, with smaller variance having larger point sizes and greater weighting in the statistical model.
